# Can Search Query Forecast successfully in China’s novel coronavirus (2019-nCov) pneumonia?

**DOI:** 10.1101/2020.02.12.20022400

**Authors:** LI Xiaoxuan, WU Qi, Lv Bbenfu

## Abstract

Recently the novel coronavirus (2019-nCov) pneumonia outbreak in China then the world, and the Number of infections and death continues to increases. Search Query performs well in forecasting the epidemics. It is still a question whether search engine data can forecast the drift and the inflexion in 2019-nCov pneumonia. Based on the Baidu Search Index, we propose three prediction models: composite Index, composite Index with filtering and suspected NCP(Novel Coronavirus Pneumonia). The result demonstrates that the predictive model of composite index with filtering performs the best while the model of suspected NCP has the highest forecast error. We further predict the out-of-the-set NCP confirmed cases and monitor that the next peak of new diagnoses will occur on February 16^th^ and 17^th^.

## Introduction

National Health Commission of PRC released a report on Unexplained Viral Pneumonia in Wuhan on Jan. 11^th^,2020. NHC shared the Gene sequence of the novel coronavirus with WHO. This virus was named as 2019-nCov officially by WHO [1]. This coronavirus has not wild spread at first. The mobility of the population has greatly increased before Lunar New Year at the end of January, and the coronavirus is contagious, the infected cases rapid growth. Up to Feb. 9^th^, the cumulative diagnosis cases is 42368 in China and 386 abroad^1^.

China has become a spotlight in the world for its current severe epidemic. China’s provinces have successively activated first-level public health emergency response since January 24. On the one hand, researchers are actively exploring effective treatment against 2019-nCov; on the other hand, they are discussing how to predict the development trend of the novel pneumonia in the future and the turning point. Existing predictions of epidemic transmitted diseases can be roughly divided into: Prediction by search engine [2][3], SNS (social networking services) [4] [5] and the data of disease prevention and control center [6] and sentinel hospitals. Because of its abundant data and its characteristics of strong periodicity and seasonal outbreaks, most of the existing studies are aimed at the predication of influenza outbreaks, which are divided into two categories: (1) nowcasting. GFT is the most representative and widely used predictive model. This model uses a range of flu keywords in Google’s search engine, and the system will automatically track and analyze the flu as long as the users enter those keywords [7]. (2) forecasting. This kind of study focuses on exploring effective epidemic predictive model [8] for the purpose of early warning of epidemic. Yang et al. [9] put forward ARGO (Auto Regression with Google search data) which uses the autoregressive and self-adjusting of GFT series to improve the prediction accuracy, and found that the prediction error is nearly three times lower than that of GFT. When the model was used to predict flu in eight Latin American countries, the accuracy of flu prediction in six of them was higher than that in time series autoregressive model and was significantly higher than that in GFT [10]. Liu, K., Srinivasan, R., & Meyers, L. A. [11] built an early flu monitoring model based network data. This model predicts seasonal flu 16.4 weeks earlier than CDC and 5 weeks earlier than Pervaiz et al [12] using Google data. In out of sample test, it was not effective in predicting 2009HINI, mainly because 2009HINI is earlier and more transmissible than seasonal flu.

However, the 2019-nCov that outbreaks in China this time is different from flu. It is a newfound airborne new virus, with no prior data, no obvious periodicity characteristics and limited data accumulation. How can we predict this disease in this case? SARS-Cov and MERS-Cov also have these predictive characteristics. In 2004, after the outbreak of SARS-Cov in China, some scholars used Gene Expression Programming (GEP) to establish the automatic Mathematical Modeling of the spread and trend of SARS in China. By simulating the epidemic in Beijing and Shanxi, it was found that the success rate of this method was 97% [13]. During the outbreak of H5N1flu in Egypt, the prediction effect and accuracy of Random Forest Model have obvious advantages over ARIMA model [14]. When the MERS-Cov propagation prediction model was established, Bayesian Belief Network was used to establish the risk assessment based on the region. The simulation of 200,000 users shows that the model gives high prediction accuracy of the region and category risk [15]. When monitoring the outbreak of H7N9 AIV in China from 2013-2017 by using weekly data of Baidu Index and Microblog Index, it was found that Baidu Index and Microblog Index always exist in a positive correlation for a certain lead time with confirmed cases during the outbreak of flu [16], which confirms the possibility of network can predict similar temporary epidemic transmitted diseases. Recently, many researches focus on the estimation of 2019-nCov by the transmission model[17-24]. By estimating the basic reproductive number *R*_0_ to release the spread of the coronavirus during the epidemic progresses. All the models have to estimate with much assumptions and the forecasting accuracy are limited. Search data can successfully predict periodicity flu outbreaks. Can novel coronavirus pneumonia in China be predicted? Can the next diffusion turning point and outbreak peak be found? These both are problems to be solved urgently.

## Materials and Method

### Data Source

With the rapid transmission of 2019-nCov recently, the data that NHS publishes data every day at regular intervals, including the number of accumulated confirmed cases, existing confirmed cases, new confirmed cases, existing suspected cases and new suspected cases. Considering the historical additivity of the number of accumulated confirmed cases, the number of existing confirmed cases is that the number of accumulated confirmed cases minus the number of cure and death cases. However, these two data are relatively small and the number of existing suspected cases has historical additivity and some overlap with the number of confirmed cases, so this paper chooses the number of new confirmed cases and new suspected cases to predict 2019-nCov^1^. NHS began to publish the daily data of new confirmed epidemic on January 17, 2020. This paper chooses the number of new confirmed cases from January 17, 2020 to February 9, 2020, totaled 24 days. The daily new suspected cases began to publish later so we choose the number of new suspected cases from January 20, 2020 to February 9, 2020, totaled 21 days. The data of the last seven days to be kept as the prediction set and the rest as the training prediction model of training set.

The main affected area of this 2019-nCov is China, and foreign confirmed cases are also related to the flow of Chinese residents, so this paper chooses the data of Baidu, the most widely used search engine in China. Baidu Index^2^ regularly published the search volume of the previous day in the form of search terms. According to the latest research, the incubation period of 2019-nCov is 7-14 days. Baidu search data of 40 days from January 1, 2020 to February 9, 2020 were chose to ensure that Baidu data has enough lead time compared with the epidemic data.

### Query Selection and Compositing

For screening of the Search Index, most of the literature adopt the search engine recommendation method, but its prediction error is large and the search terms recommended have too much useless information and noise. By considering that this disease has a long incubation period and the symptoms of this disease are relatively defined in the clinical cases so far, it indicated that the fever accounted for 87.9% and cough accounted for 67.7% of the confirmed cases [25]. Starting with the most common “fever” and “cough”, this paper uses the demand map in Baidu search to select the relevant search terms that users demand the most in the change of search behavior before and after search terms. In addition, with the increasing understanding of the disease, the common search terms for the disease “the novel coronavirus pneumonia” and “Wuhan pneumonia” were added, and the search volume of the rest of common search terms almost coincides with the outbreak of the epidemic, which could be initially judged as the event-based search noise and could not be used as the search term for the outbreak prediction. After screening the search terms, and removing the unrecorded terms, this paper retains 16 search terms in total. Using Pearson Correlation Coefficient Method, the most commonly used in network data prediction, confirms the correlation and the lead time between search terms and new confirmed sequence. By calculating the correlation coefficient of each word from lead 16 periods to 0 period and new confirmed cases, it is founded that all search terms have a high linear dependence with the new confirmed sequence, which shows that the selected search terms have good prediction ability. Most of the search terms are 10 days earlier than the new confirmed sequence while new suspected sequence is 4 earlier than new confirmed sequence (correlation coefficient is 0.8914). Table 1 shows the maximum correlation coefficient between search terms and new confirmed sequence.

**Table1.** The maximum correlation coefficient with NCP.

| Search Query | MCC | Search Query | MCC |
| --- | --- | --- | --- |
| 咳嗽<br>(cough) | 0.9164 | 干咳<br>(dry cough) | 0.8711 |
| 呼吸困难<br>(dyspnea) | 0.9129 | 胸闷<br>(chest congestion) | 0.9133 |
| 发热<br>(pyrexia) | 0.8974 | 发烧<br>(fever) | 0.9201 |
| 发烧温度<br>(fever temperature) | 0.9235 | 多少度算发烧<br>(how many degrees is a fever) | 0.9305 |
| 低烧<br>(low fever) | 0.9101 | 感冒<br>(cold) | 0.8749 |
| 发烧的症状<br>(symptoms of fever) | 0.9578 | 乏力<br>(exhaustion) | 0.8919 |
| 四肢无力<br>(limb weakness) | 0.8758 | 腹泻<br>(diarrhea) | 0.8375 |
| 新型冠状病毒<br>(new coronavirus) | 0.9193 | 武汉肺炎<br>(Wuhan pneumonia) | 0.8911 |

\*MCC is the abbreviation for Maximum Correlation Coefficient.

The correlation coefficient among the 16 search terms and new confirmed cases are all above 0.8 and leading, which indicates that these search terms have certain prediction ability for future epidemic. Based on the search terms, all terms are combined into search composite index 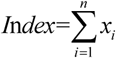 by simple linear sum, and *x*_*i*_ is the search volume for each sequence. Recalculating the correlation coefficient after combination, it is found that the highest correlation coefficient is lead new confirmed 10 times, 0.9248. Fig. 1 shows the sequence diagram between search composite index and new confirmed cases.

**Figure1.**
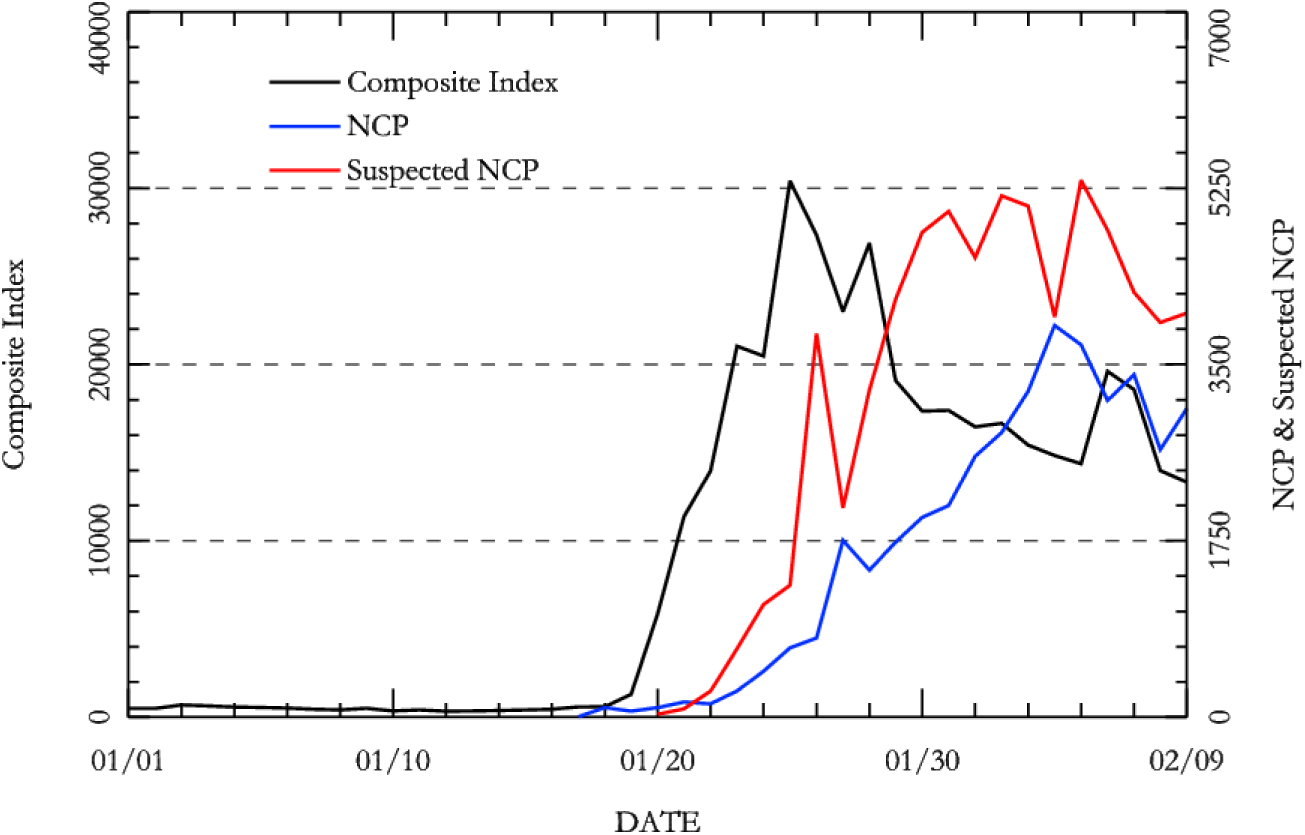
NCP, Suspected NCP and Composite Index.

### Model

On the basis of determining the time correlation between the search composite *Index* and new suspected cases corresponding to the new confirmed cases, the training set can be used to train and fit the prediction model, and the prediction set can be used to verify prediction model based on the training. Li et al. [26] presented the effect of noise interference on the prediction in network search data and search data prediction ability will improve significantly after eliminating the high frequency noise on search data. In physics, Fourier transform, a common signal analysis method, is used to reveal the intrinsic relation between the time function and the spectral function in the signal and to extract the signal of the highest frequency sequence by using all the temporal information of the signal to deal with the high frequency noise. Table 2 shows the filtering result. The search composite index after filtering is smoother than original sequence.

**Table2.** Statistical Results for Models.

| Variable | Coefficient | Std. Error | t-statistic | Prob. | R-squared | DW stat |
| --- | --- | --- | --- | --- | --- | --- |
| $Index_{t-10}$ | 0.116 | 0.019 | 5.953 | <0.001 | 0.904 | 1.911 |
| $Index\_filtering_{t-10}$ | 0.106 | 0.022 | 4.689 | 0.0005 | 0.936 | 1.824 |
| $NCPS_{t-4}$ | 0.611 | 0.075 | 8.142 | <0.001 | 0.815 | 1.854 |
|  | t-stat | 1% | 5% | 10% | Prob. | result |
| Residual stationary | -3.256 | -2.717 | -1.964 | -1.606 | 0.003 | stationary |
|  | -3.553 | -2.717 | -1.964 | -1.606 | 0.001 | stationary |
|  | -3.112 | -2.717 | -1.964 | -1.606 | 0.004 | stationary |

Based on the above, three 2019-nCov prediction models are established respectively in this paper: the prediction model based on the search composite index *Index*, the prediction model based on the search composite index for filtering noise reduction *Index* _ *filtering* and prediction model based on new suspected cases:

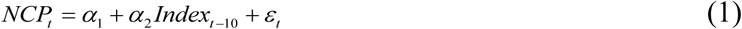

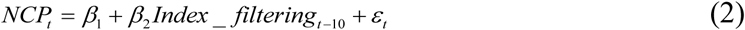

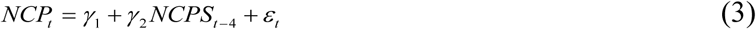

To be specific, *NCP*_*t*_ denotes the new confirmed cases of 2019-nCov in t period, *Index*_*t* −10_ denotes search composite index of the first 10 periods, *Index* _ *filtering*_*t* −10_ denotes the search composite index of the first 10 periods after filtering and *NCPS*_*t* −4_ denotes the first 4 periods of new suspected cases of 2019-nCov. Search composite index shows the results of netizen’s self-examination of the symptoms of viral pneumonia, which can be used to explain the spread and diffusion of viral pneumonia in the future. The number of daily new suspected case is the people who have had related symptoms but not have been confirmed with the virus through actual detection. The suspected cases include people who are actually infected with the virus but cannot be detected temporarily because of current medicine, and have a strong correlation and explanation with the number of the future confirmed cases. First, the sequence was tested for ADF and the results show that all the original sequences were not steady, but there is no unit root in the first difference sequence at 5% significance level. First difference sequence is a stationary sequence, so Granger Non-Causal Test and ARMAX model can be supported. Table 2 shows the model results by training set data.

From the above model fitting results, it can be seen that the prediction model based on filtering noise reduction has the best fitting effect among the three prediction model, followed by the prediction model based on search composite index. Major variables are all significant in 1%, which indicates that search composite index *Index*, search composite index after filtering and noise reduction *Index* _ *filtering* and *NCPS* are all have some explanatory and prediction ability to the new confirmed cases in the future.

### Epidemic Forecast

Based on the above training models, this paper predicts the new confirmed cases of 2019-nCov from February 3 to 9, 2020 in prediction set and evaluates the prediction effect of the three prediction models. Among many evaluation indexes for prediction effect, this paper chooses the common prediction error index-MAPE and RMSE. The predictive values of China 2019-nCov in the next seven days in prediction set are shown in Table 3.

**Table3.** Predicted Values and Error.

| Date | Model | 2/3 | 2/4 | 2/5 | 2/6 | 2/7 | 2/8 | 2/9 |
| --- | --- | --- | --- | --- | --- | --- | --- | --- |
| **Actual Value** |  | 3233 | 3886 | 3694 | 3143 | 3399 | 2656 | 3062 |
| **Predicted Value** | $Index_{t-10}$ | 2971 | 4120 | 3766 | 3263 | 3713 | 2809 | 2610 |
| | $Index\_filtering_{t-10}$ | 3235 | 3615 | 3632 | 3440 | 3221 | 2895 | 2987 |
| | $NCPS_{t-4}$ | 3268 | 3386 | 3126 | 3473 | 3416 | 2791 | 3561 |
| **Absolute Error** | $Index_{t-10}$ | 262 | 234 | 72 | 120 | 314 | 153 | 452 |
| | $Index\_filtering_{t-10}$ | 2 | 271 | 62 | 297 | 178 | 239 | 75 |
| | $NCPS_{t-4}$ | 35 | 500 | 568 | 330 | 17 | 135 | 499 |
| MAPE | $Index_{t-10}$ | 7.09% | | | | | | |
| | $Index\_filtering_{t-10}$ | 4.98% | | | | | | |
| | $NCPS_{t-4}$ | 8.82% | | | | | | |
| RMSE | $Index_{t-10}$ | 258.97 | | | | | | |
| | $Index\_filtering_{t-10}$ | 192.71 | | | | | | |
| | $NCPS_{t-4}$ | 368.51 | | | | | | |

It can be seen from the prediction results during the above prediction period that among the three prediction models, the model with the smallest prediction error is search composite index after filtering and noise reduction, whose MAPE and RMSE are 4.98% and 192.71 respectively. The second model is search composite index, whose MAPE and RMSE are 7.09% and 258.97 respectively. By contrast, the prediction accuracy of predicting new confirmed cases by the new suspected cases of the first four periods is the worst and the prediction lead time is shorter. Fig. 3 shows the 2019-nCov original value and prediction value during the prediction period.

**Figure2.**
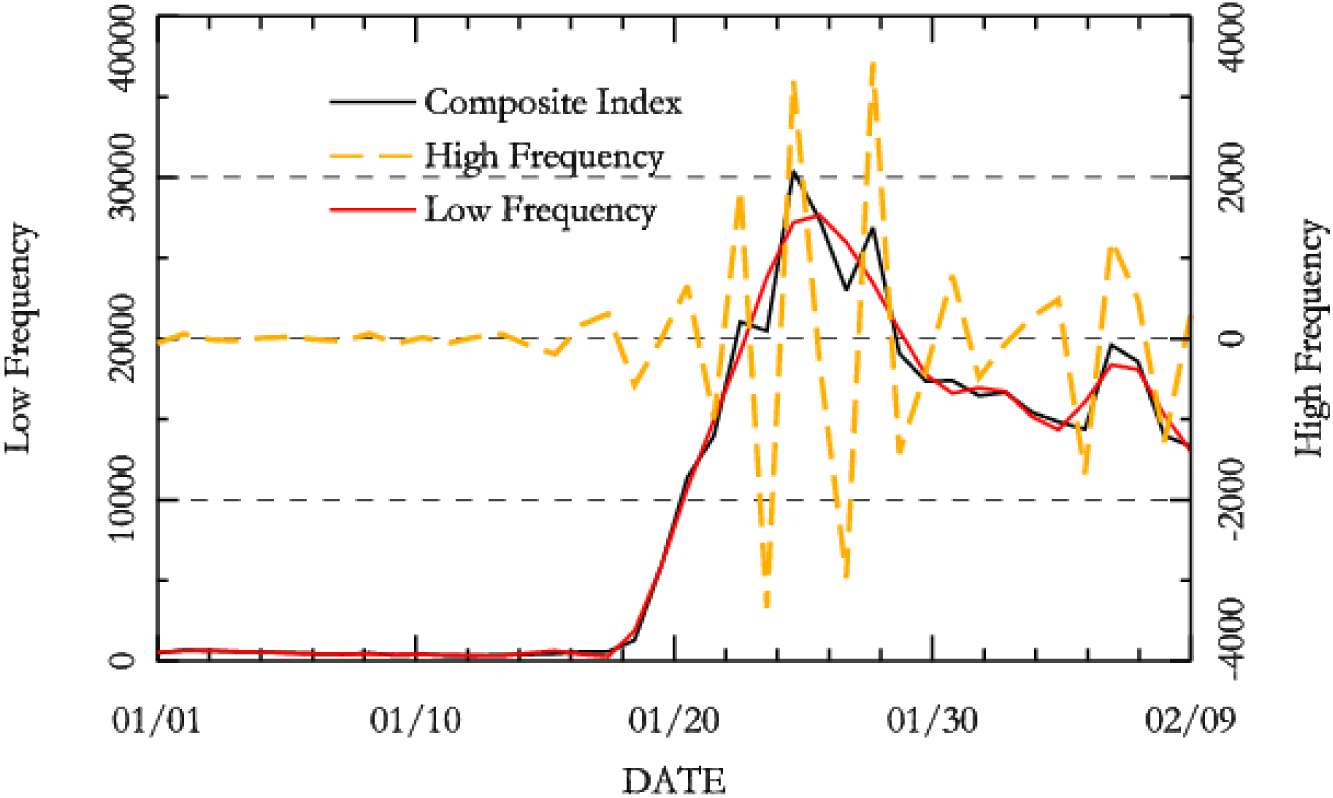
Fourier transform filtering.

**Figure3.**
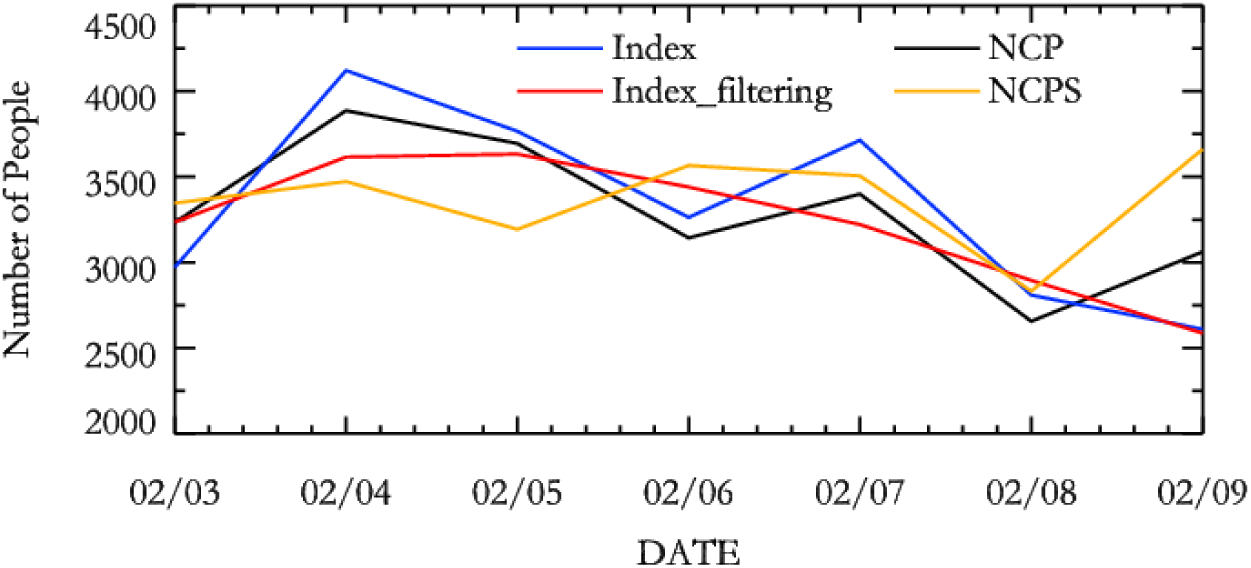
NCP and prediction based on models.

**Figure4.**
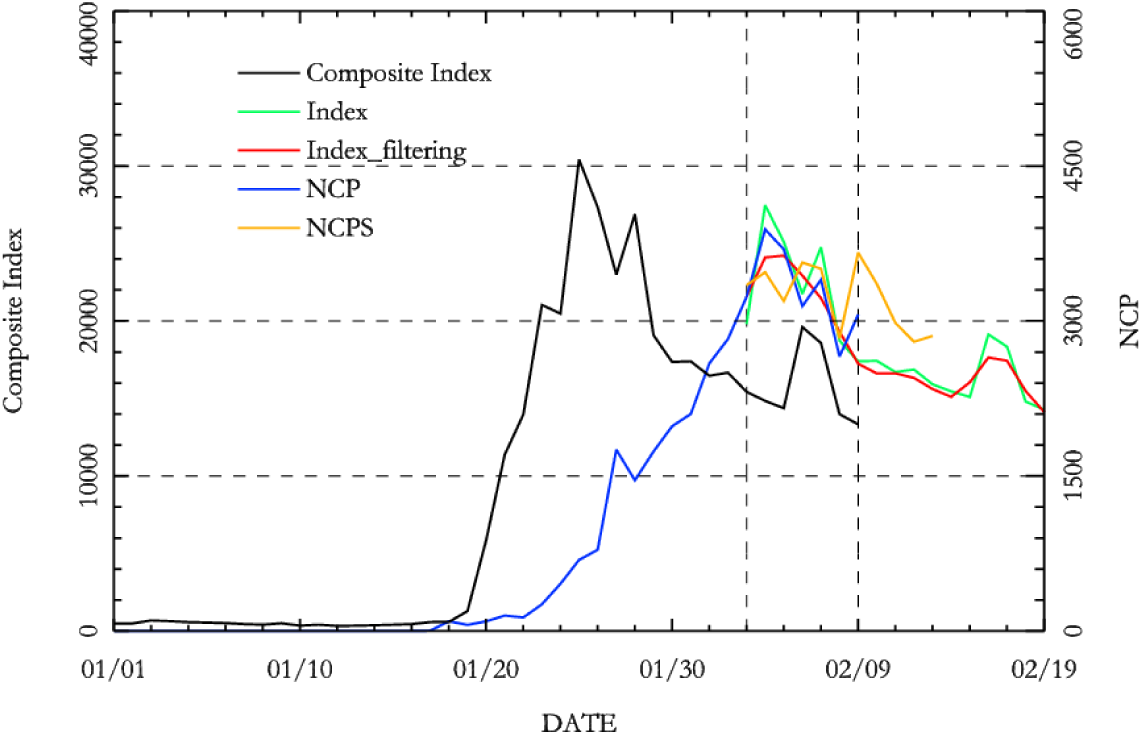
Further forecasting of NCP.

It can be concluded from the absolute error that the prediction accuracy of search composite index after filtering and noise reduction is the highest but the overall prediction trend is flat due to filter-out of high frequency noise. Therefore, the peak change can’t be obtained in a short period. However, this defect can be well solved by search composite index *Index*. It can be seen from prediction value that the prediction value of search composite index at every turning point is overestimated or underestimated but the prediction accuracy of the short-term fluctuation is high. According to the above analysis, the prediction model of search composite index after filtering and noise reduction can be used to observe the epidemic trend in the long future period and the prediction model based on search composite index can be used to monitor the epidemic fluctuation in the short period.

At the end of this paper, the change trend of the epidemic in the next 10 days from February 9 to 18 is further obtained according to the prediction model after evaluating the prediction effect of the prediction model by using the prediction set, and the results are shown in Table 4.

It can be seen from the figure that the search composite index and index after filtering and noise reduction can be used for predicting the 2019-nCov in the coming 10 days and the new suspected cases can be used for predicting the 2019-nCov in the coming four days. The search composite index reached the peak on January 23, 25 and 28 respectively, which basically coincide with the three small peaks of new confirmed cases on February 4, 6 and 9 respectively. It embodies the apriority of the index. The index has been declining thereafter but met another search peak on February 6 and 7 respectively. It means the new confirmed cases of 2019-nCov in the coming period would also meet a small peak after continuous decline. The result of the prediction model displays the peak is expected to appear on February 16 and 17 respectively. The new suspected cases can be used for predicting the number of new confirmed cases in the coming 4 days only. The prediction result on February 10-13 is basically in consistency with the other two models, which means the third small peak will appear on February 9 and then the case will decline for a short period.

## Discussion

Under the current rigid status of 2019-nCov in the world, particularly in China, this paper uses Baidu search index to predict the viral pneumonia and brings forth three prediction models: (1) Prediction model based on Baidu search composite index; (2) Prediction model based on search composite index after filtering and noise reduction; (3) Prediction model based on the suspected cases increased every day. The three models can all be used to predict the new confirmed persons suffering 2019-nCov in the coming period. To be specific, the former two prediction models can be used to predict the new confirmed persons in the coming ten days while the third model can be used to predict the new confirmed persons in the coming 4 days. It can be concluded from the model prediction effect that search composite index based on filtering and noise reduction has the highest prediction accuracy for new confirmed persons but it is not sensitive to short-term fluctuation; the prediction accuracy based on search composite index is sensitive to prediction of peak and valley although its prediction error is larger than Model (2). Prediction accuracy based on new suspected cases is the worst. Therefore, Model (2) can be used to predict the epidemic change trend in the future and can also be used to monitor short-term epidemic fluctuation by working together with Model (1). By further predicting the new confirmed cases in the coming ten days beyond the prediction period at last, this paper predicts that the new confirmed cases of 2019-nCov will meet a small peak on February 16 and 17 respectively, after which, it will decline in an oscillated manner.

The prediction capacity of the current’s prediction model is limited for the 2019-nCov, an emergent infectious disease. This paper just brings forth a prediction idea and the prediction precision needs further improvement, which also implies the arduous task for epidemic forecast.

## Data Availability

the new confirmed cases and the new suspected cases is availabe from National Health Commission of China.
The Baidu search engines data is available from the Baidu Website.

http://www.nhc.gov.cn/xcs/xxgzbd/gzbd_index.shtml

http://index.baidu.com/v2/index.html#/

## Ethics approval and consent to participate

The ethical approval or individual consent was not applicable

## Footnotes

1 http://www.nhc.gov.cn/xcs/xxgzbd/gzbd_index.shtml

## References

[1] Hui D S, I Azhar E, Madani T A, et al. The continuing 2019-nCoV epidemic threat of novel coronaviruses to global health—The latest 2019 novel coronavirus outbreak in Wuhan, China[J]. International Journal of Infectious Diseases, 2020, 91: 264–266.

[2] Ning, Shaoyang, Shihao Yang, and S. C. Kou. “Accurate regional influenza epidemics tracking using Internet search data.” Scientific reports 9.1 (2019): 1–8.

[3] Santillana, Mauricio, et al. “Combining search, social media, and traditional data sources to improve influenza surveillance.” PLoS computational biology 11.10 (2015):1–15.

[4] Lu, Fred S., et al. “Improved state-level influenza nowcasting in the United States leveraging Internet-based data and network approaches.” Nature communications 10.1 (2019): 1–10.

[5] Xue, Hongxin, et al. “Regional level influenza study based on Twitter and machine learning method.” PloS one 14.4 (2019):1–23.

[6] Basile, Luca, et al. “Real-time predictive seasonal influenza model in Catalonia, Spain.” PloS one 13.3 (2018):1–15.

[7] Ginsberg, Jeremy, et al. “Detecting influenza epidemics using search engine query data.” Nature 457.7232 (2009): 1012–1014.

[8] Reich, Nicholas G., et al. “A collaborative multiyear, multimodel assessment of seasonal influenza forecasting in the United States.” Proceedings of the National Academy of Sciences 116.8 (2019): 3146–3154.

[9] Yang, Shihao, Mauricio Santillana, and Samuel C. Kou. “Accurate estimation of influenza epidemics using Google search data via ARGO.” Proceedings of the National Academy of Sciences 112.47 (2015): 14473–14478.

[10] Clemente, Leonardo, Fred Lu, and Mauricio Santillana. “Improved Real-Time Influenza Surveillance: Using Internet Search Data in Eight Latin American Countries.” JMIR public health and surveillance 5.2 (2019): e12214.

[11] Liu, Kai, Ravi Srinivasan, and Lauren Ancel Meyers. “Early Detection of Influenza outbreaks in the United States.” arXiv preprint 1903.01048 (2019).

[12] Pervaiz F, Pervaiz M, Rehman NA, Saif U. FluBreaks: early epidemic detection from Google flu trends. Journal of medical internet research. 2012 Sep;14(5).

[13] Huang L, Xiao S. Application of SARS epidemic situation analysis and forecast based on gene expression programming[J]. Computer Engineering, 2007, 33(4): 45–48.

[14] Kane, Michael J., et al. “Comparison of ARIMA and Random Forest time series models for prediction of avian influenza H5N1 outbreaks.” BMC bioinformatics 15.1 (2014): 276–285.

[15] Sandhu, Rajinder, Sandeep K. Sood, and Gurpreet Kaur. “An intelligent system for predicting and preventing MERS-CoV infection outbreak.” The Journal of Supercomputing 72.8 (2016): 3033–3056.

[16] Chen Y, Zhang Y, Xu Z, et al. Avian Influenza A (H7N9) and related Internet search query data in China[J]. Scientific reports, 2019, 9(1): 1–9.

[17] Wang, Huwen and Wang, Zezhou and Dong, Yinqiao and Chang, Ruijie and Xu, Chen and Yu, Xiaoyue and Zhang, Shuxian and Tsamlag, Lhakpa and Shang, Meili and Huang, Jinyan and Wang, Ying and Xu, Gang and Shen, Tian and Zhang, Xinxin and Cai, Yong, Estimating the Number of 2019 Novel Coronavirus Cases in Chinese Mainland (1/29/2020). Available at SSRN: https://ssrn.com/abstract=3529449

[18] Imai N, Cori A, Dorigatti I, et al. Report 3: Transmissibility of 2019-nCoV[J]. Reference Source, 2020.

[19] Liu T, Hu J, Kang M, et al. Transmission dynamics of 2019 novel coronavirus (2019-nCoV)[J]. 2020.

[20] Read J M, Bridgen J R E, Cummings D A T, et al. Novel coronavirus 2019-nCoV: early estimation of epidemiological parameters and epidemic predictions[J]. medRxiv, 2020.

[22] Riou J Y, Althaus C. Pattern of early human-to-human transmission of Wuhan 2019 novel coronavirus (2019-nCoV), December 2019 to January 2020[J]. Eurosurveillance, 2020, 25(4): pii= 2000058.

[23] Zhao S, Lin Q, Ran J, et al. Preliminary estimation of the basic reproduction number of novel coronavirus (2019-nCoV) in China, from 2019 to 2020: A data-driven analysis in the early phase of the outbreak[J]. International Journal of Infectious Diseases, 2020.

[24] Park S W, Champredon D, Earn D J D, et al. Reconciling early-outbreak preliminary estimates of the basic reproductive number and its uncertainty: a new framework and applications to the novel coronavirus (2019-nCoV) outbreak[J]. medRxiv, 2020.

[25] Kang M, Wu J, Ma W, et al. Evidence and characteristics of human-to-human transmission of 2019-nCoV[J]. medRxiv, 2020.

[26] Xiaoxuan L, Qi W, Geng P, et al. Tourism forecasting by search engine data with noise-processing[J]. African Journal of Business Management, 2016, 10(6): 114–130.

